# Carotid endarterectomy is associated with lower rates of reintervention compared to carotid stenting

**DOI:** 10.1101/2025.01.13.25320499

**Authors:** Shaunak S. Adkar, Elizabeth L. George, Xinyan Zheng, Sabina M. Sorondo, Arash Fereydooni, Shernaz Dossabhoy, Jordan R. Stern

## Abstract

**Objectives:** Perioperative outcomes for carotid endarterectomy (CEA) and transfemoral carotid artery stenting (TFCAS) have been well studied. Less is known about the durability and reintervention rates of each, particularly in the era of transcarotid artery revascularization (TCAR). We sought to compare real-world rates of ipsilateral reintervention, stroke, and death in patients undergoing CEA, TFCAS, and TCAR.

**Methods:** The Vascular Quality Initiative (VQI) was matched to Medicare claims via the Vascular Implant Surveillance and Implantation Network (VISION) database to identify patients who had primary carotid revascularization from December 2016 to December 2019 in a n observational cohort study. The primary outcome was ipsilateral reintervention; secondary outcomes included stroke and mortality. After 1:1 greedy matching using propensity scores, patients who underwent CEA and carotid artery stenting (CAS) via either transcarotid or transfemoral approach were compared using time-dependent Cox regression models. A separate propensity matched analysis was then performed to compare TFCAS and TCAR. Kaplan-Meier curves were compared using log rank tests.

**Results:** After propensity matching (N=27,944 patients), we compared 4705 patients in each group. Risk of re-intervention was increased within 6 months for CAS (HR: 1.97; 95% CI: 1.11-3.50; p<0.05), but not beyond 6 months (HR: 1.08; 95% CI: 0.62-1.89; p=0.79). The incidence of stroke prior to discharge was increased in patients undergoing CAS (5.4% v. 1.0%; p<0.0001) and mortality hazard with CAS was increased both within 6 months (HR:1.69; 95% CI: 1.38- 2.07; p<0.0001) and beyond 6 months (HR: 1.52; 95% CI: 1.27-1.81; p<0.0001). When comparing TFCAS and TCAR (n=2115 per group), there was a significantly increased risk of re- intervention for TFCAS beyond 6 months (HR: 2.31, 95% CI: 1.05-5.11, p <0.05).

**Conclusions:** CEA portends a lower risk of reintervention than CAS, particularly within the first 6 months after revascularization. On subgroup analysis of stenting modalities, TCAR had a lower hazard of longer-term reintervention compared to TFCAS.

## Introduction

Extracranial carotid artery stenosis accounts for 10-15% of all ischemic strokes.^1^ High quality evidence from randomized controlled trials (RCTs) and observational studies supports the use of carotid endarterectomy (CEA) as a revascularization strategy for carotid artery stenosis to prevent the risk of stroke.^2–6^ Patients with prohibitive high-risk physiologic or anatomic factors, or clinical history (e.g. neck radiation) often undergo carotid artery stenting (CAS).^7^ Classically, this has been accomplished via a transfemoral approach (TFCAS); however, in recent years transcarotid artery revascularization (TCAR) with flow reversal has emerged as an option.^8–11^ Perioperative and short-term outcomes following these procedures have been well studied. The CREST trial demonstrated that patients undergoing CEA have a reduced risk of perioperative stroke compared to patients undergoing TFCAS.^6^ In addition, the ROADSTER trial reported a 30-day stroke rate at 1.4% for TCAR, on par with stroke incidence following CEA.^12^ In a relatively small cohort, the ongoing ROADSTER2 trial reported an extremely low incidence of stroke (0.4%) over a 1-year post-operative period.^13^

The use of TCAR as a revascularization strategy has grown concomitant with perioperative data supporting its use; however, intermediate and long-term outcomes compared to CEA and CAS remain unclear. In particular, the rate of reintervention of TCAR remains to be defined. We therefore sought to examine mid- to long-term rates of reintervention for CEA compared to transfemoral and transcarotid stenting modalities using the Vascular Quality Initiative (VQI) dataset linked to Medicare claims data to obtain longer than 21-month follow- up.

## Methods

The VQI registries are a validated clinical database that are comprised of demographic, procedural, and outcomes data of patients undergoing vascular procedures from over 1000 centers and 18 regions in the United States, Canada, and Singapore. The Vascular Implant Surveillance and Implantation Network (VISION) is a collaborative effort between the VQI and the Medical Device Epidemiology Network (MDEpiNet) that links data from VQI registries to Medicare claims data. VISION provides long-term outcomes related to mortality, adverse events, reintervention, readmission, imaging, and cost that are not provided in the VQI registries.

For these analyses, we leveraged data from the VQI carotid stent and endarterectomy registries linked to Medicare claims data using VISION to identify patients with primary carotid artery revascularization from 2016-2019 with 36-month follow-up data available in VISION. This yielded 121,197 patients. We excluded patients who had both CEA and CAS as the index procedure, CAS approach other than femoral, or trans-carotid/open carotid, and those with no Medicare part A and part B enrollment at the time of the index operation. We employed 1:1 greedy propensity score matching to control baseline differences in demographics. Comparisons of patient and procedure related characteristics were performed with Pearson χ^2^ test for categorical variables and Wilcoxon rank sum test for continuous variables.

The primary outcome for this analysis was ipsilateral reintervention within the 36-month study period. To determine who had ipsilateral reinterventions, we identified patients in the VISION dataset who had ipsilateral restenosis along with an ICD10-PCS or CPT code for CAS or CEA following the index operation. Secondary endpoints included ipsilateral cerebrovascular accident, stroke and transient ischemic attack (TIA), and mortality over the study period. We generated unadjusted Kaplan-Meier curves and calculated the hazard ratio of the primary and secondary outcomes within 6 months of the index operation and from 6 months to 36 months using time-dependent Cox regression models. We performed a post-hoc subgroup analysis of patients undergoing primary carotid stenting, separately comparing TCAR and TFCAS to each other and then to CEA, again using greedy propensity matched cohorts and time-dependent Cox regression. To identify patient and procedure related variables that confer risk of reintervention, we used multivariable logistic regression. This study was approved by the Stanford University Institutional Review Board (IRB-57282) and follows STROBE guidelines for observational studies.

## Results

Using the 2016-2019 VQI-VISION dataset, we identified 27,913 patients who had undergone CEA (n=21,312) or CAS (n=6,688) and met inclusion/exclusion criteria (Figure 1). After 1:1 greedy propensity score matching, we compared 4,705 patients who had undergone either CEA or CAS. Patient and procedure-related characteristics before and after propensity matching are shown in Table I. After propensity-matching, patients undergoing CAS were more likely to have coronary artery disease (CAD) (47.2% vs. 43.7%, p < 0.001), be on anticoagulation (14.2% vs. 12.3%, p < 0.01), and have a history of radiation (4.1% vs. 3.2%, p < 0.05), but were otherwise similar.

**Figure 1:**
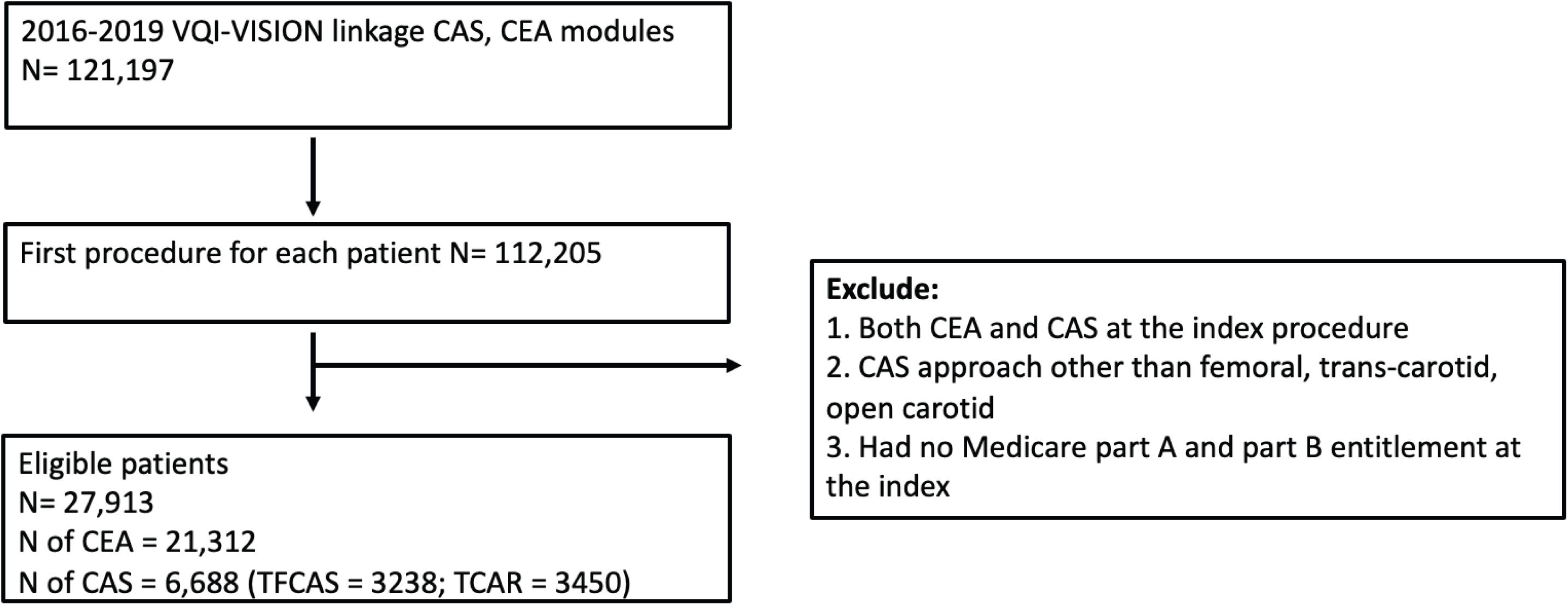
Cohort Generation. Flow diagram describing inclusion and exclusion criteria for CEA and CAS cohort generation. CEA Carotid Endarterectomy. CAS Carotid Artery Stenting.

**Table I:**
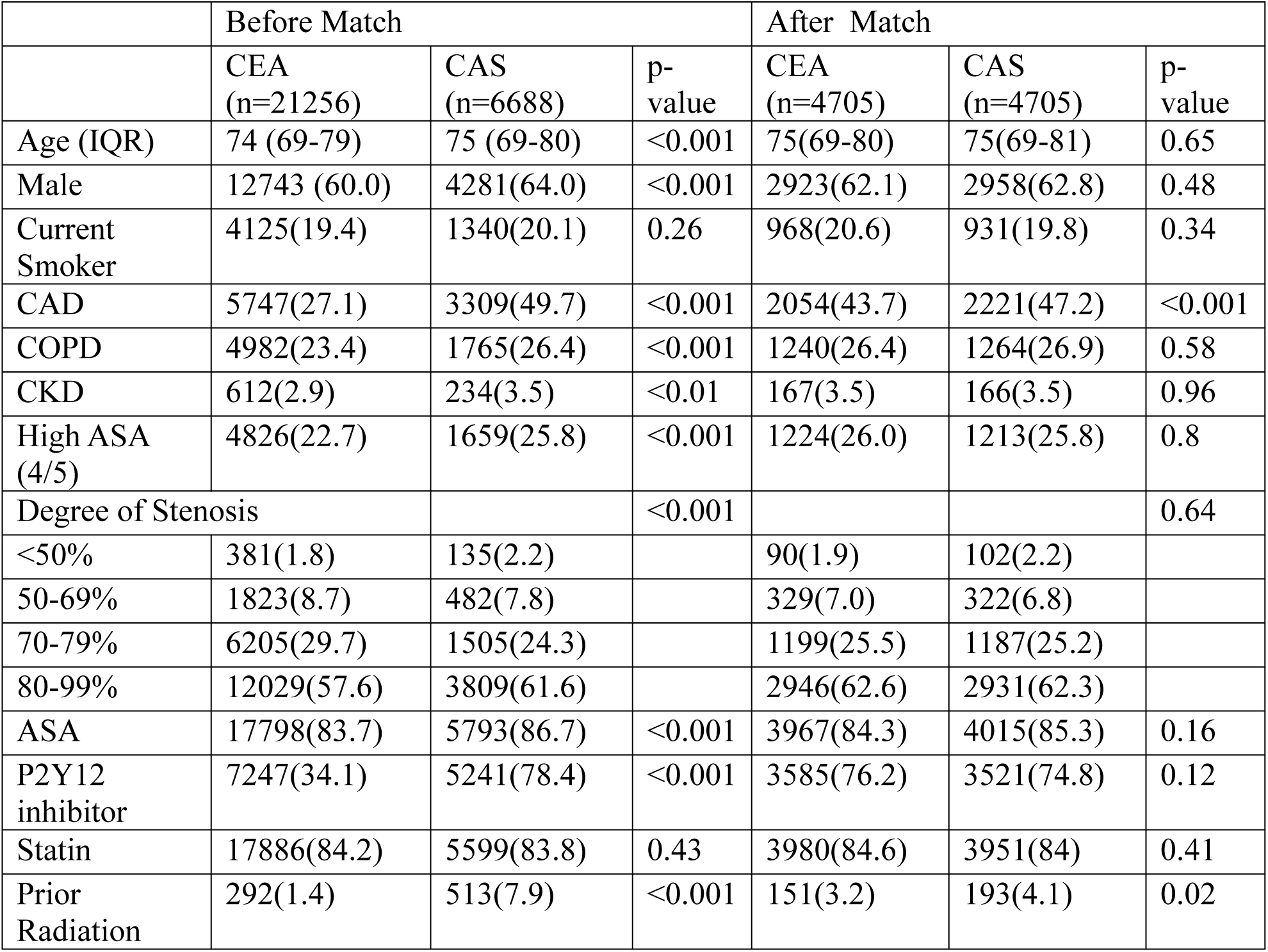
Patient-related characteristics of CEA and CAS cohorts before and after matching.

|  | Before Match |  |  | After Match |  |  |
| --- | --- | --- | --- | --- | --- | --- |
|  | CEA<br>(n=21256) | CAS<br>(n=6688) | p-value | CEA<br>(n=4705) | CAS<br>(n=4705) | p-value |
| Age (IQR) | 74 (69-79) | 75 (69-80) | <0.001 | 75(69-80) | 75(69-81) | 0.65 |
| Male | 12743 (60.0) | 4281(64.0) | <0.001 | 2923(62.1) | 2958(62.8) | 0.48 |
| Current<br>Smoker | 4125(19.4) | 1340(20.1) | 0.26 | 968(20.6) | 931(19.8) | 0.34 |
| CAD | 5747(27.1) | 3309(49.7) | <0.001 | 2054(43.7) | 2221(47.2) | <0.001 |
| COPD | 4982(23.4) | 1765(26.4) | <0.001 | 1240(26.4) | 1264(26.9) | 0.58 |
| CKD | 612(2.9) | 234(3.5) | <0.01 | 167(3.5) | 166(3.5) | 0.96 |
| High ASA<br>(4/5) | 4826(22.7) | 1659(25.8) | <0.001 | 1224(26.0) | 1213(25.8) | 0.8 |
| Degree of Stenosis |  |  | <0.001 |  |  | 0.64 |
| <50% | 381(1.8) | 135(2.2) |  | 90(1.9) | 102(2.2) |  |
| 50-69% | 1823(8.7) | 482(7.8) |  | 329(7.0) | 322(6.8) |  |
| 70-79% | 6205(29.7) | 1505(24.3) |  | 1199(25.5) | 1187(25.2) |  |
| 80-99% | 12029(57.6) | 3809(61.6) |  | 2946(62.6) | 2931(62.3) |  |
| ASA | 17798(83.7) | 5793(86.7) | <0.001 | 3967(84.3) | 4015(85.3) | 0.16 |
| P2Y12<br>inhibitor | 7247(34.1) | 5241(78.4) | <0.001 | 3585(76.2) | 3521(74.8) | 0.12 |
| Statin | 17886(84.2) | 5599(83.8) | 0.43 | 3980(84.6) | 3951(84) | 0.41 |
| Prior<br>Radiation | 292(1.4) | 513(7.9) | <0.001 | 151(3.2) | 193(4.1) | 0.02 |

We first compared outcomes following CEA compared to all CAS modalities. Patients undergoing CAS had an increased risk of in-hospital stroke (5.4% vs. 1.0%, p<0.0001). Over a 36-month follow-up period, we observed an increased incidence of stroke and mortality in patients undergoing CAS compared to CEA, consistent with prior studies (Figure 2). For CAS there was an increased stroke risk within the first six months of the procedure (HR 2.91; 95% CI:2.42-3.48; p < 0.0001), but not beyond six months (HR 1.3; 95% CI: 0.98-1.72). Risk of mortality was increased in patients undergoing CAS vs CEA both within six months (HR 1.69; 95% CI:1.38-2.07; p < 0.0001) and beyond six months (HR 1.52; 95% CI: 1.27-1.81; p < 0.0001) of the index procedure. The risk of reintervention following CEA versus CAS was not significantly different during the overall study period (Figure 1c). However, patients undergoing CAS had a significantly increased risk of reintervention within 6 months of their index procedure (HR 1.97; 95% CI: 1.11-3.50; p < 0.001) that was not sustained beyond 6 months (HR 1.08; 95% CI: 0.62-1.89).

**Figure 2:**
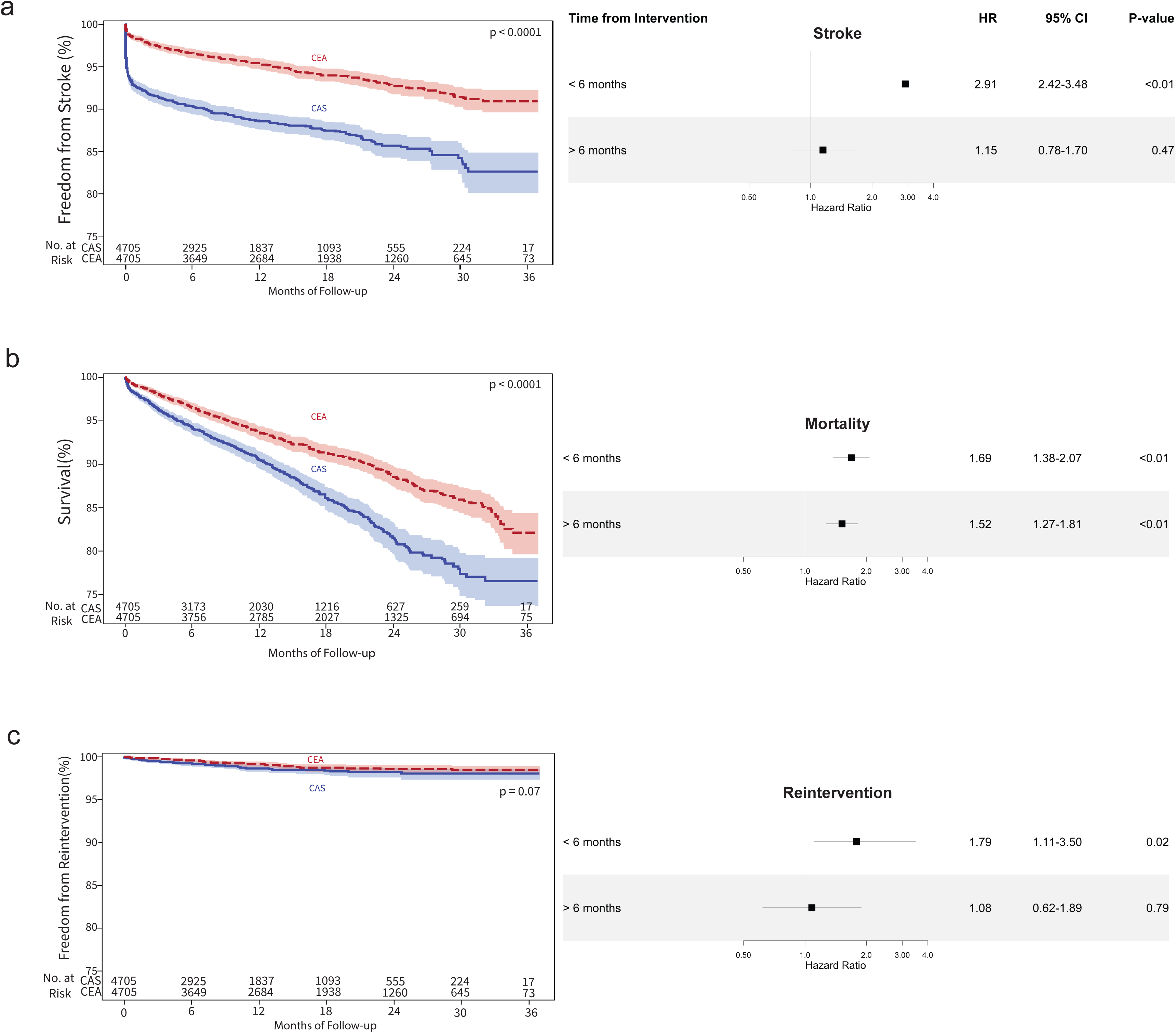
Kaplan-Meier curves and forest plots depicting freedom from a) stroke, b) mortality, and c) reintervention amongst patients undergoing CEA and CAS within the 36-month study period. HR Hazard Ratio; CI Confidence Interval

On post-hoc analysis stratifying CAS by TFCAS and TCAR (n=2115 in each arm), patients undergoing TFCAS had significantly increased risk of reintervention compared to TCAR (Table II). In addition to increased risk of reintervention, patients undergoing TFCAS also had increased risk of mortality and stroke over the course of 36 months of follow-up (Figure 3a-c). In time-dependent stratified analysis the overall increased risk of reintervention for TFCAS can be attributed to an increased risk of reintervention beyond six months of the procedure (HR: 2.31; 95% CI: 1.05-5.11, p < 0.05) (Figure 3c).

**Figure 3:**
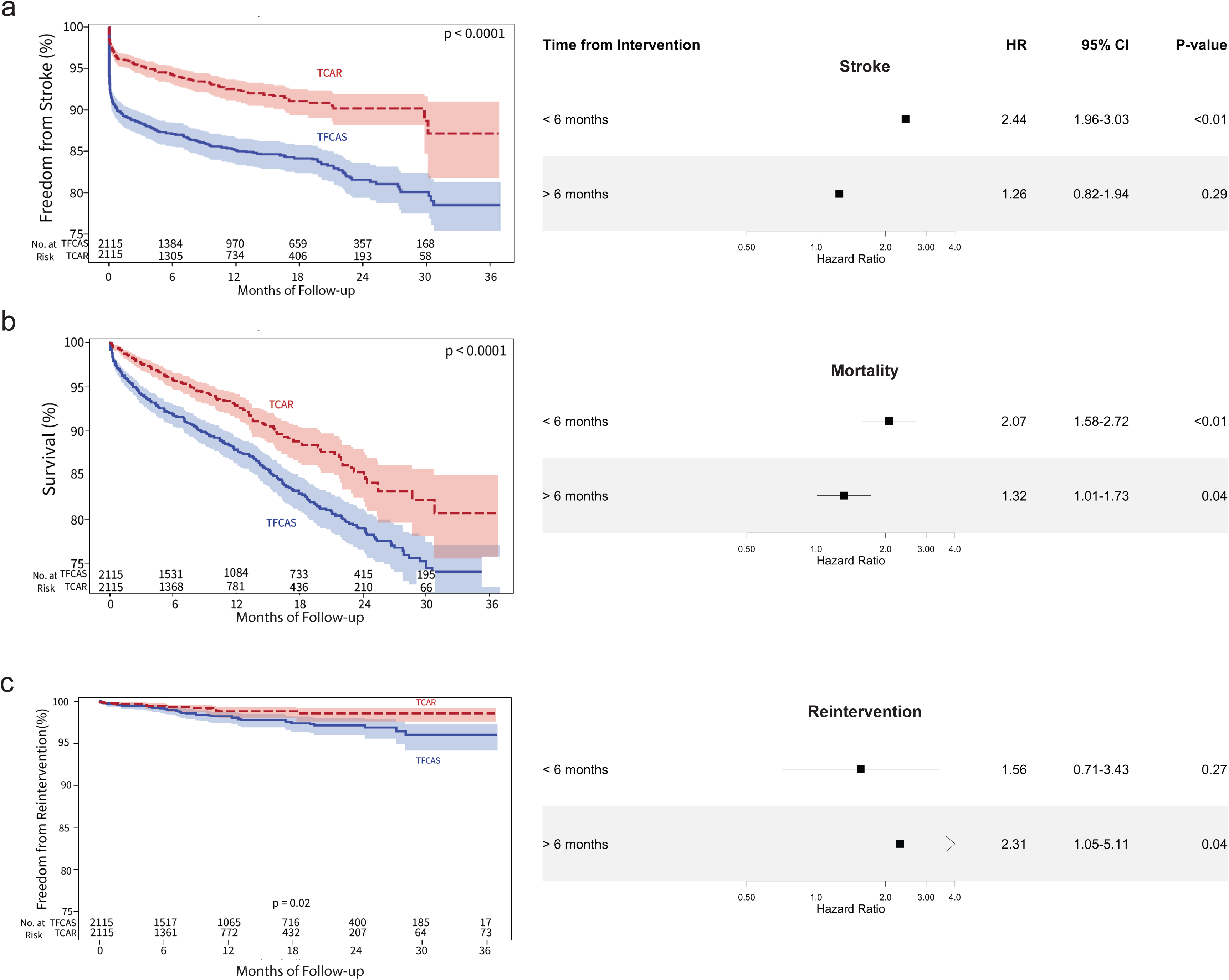
Kaplan-Meier curves and forest plots depicting freedom from a) stroke, b) mortality, and c) reintervention amongst patients undergoing TCAR and TFCAS within the 36-month study period. HR Hazard Ratio; CI Confidence Interval

**Table II:** Patient-related characteristics of TFCAS and TCAR cohorts before and after propensity score matching.

|  | Before match |  |  | After match |  |  |
| --- | --- | --- | --- | --- | --- | --- |
|  | TCAR<br>N=3450 | TFCAS<br>N=3238 | P | TCAR<br>N=2115 | TFCAS<br>N=2115 | P |
| Age: median (IQR) | 76(71-81) | 73(68-79) | <.0001 | 74(69-80) | 74(69-80) | 0.85 |
| Male | 2177(63.1%) | 2104(65.0%) | 0.11 | 1369(64.7%) | 1379(65.2%) | 0.74 |
| Smoke |  |  | <0.001 |  |  | 0.60 |
| Never | 1032(29.9%) | 1007(31.2%) |  | 624(29.5%) | 645(30.5%) |  |
| Prior | 1836(53.2%) | 1466(45.4%) |  | 1055(49.9%) | 1022(48.3%) |  |
| Current | 582(16.9%) | 758(23.5%) | <0.001 | 436(20.6%) | 448(21.2%) | 0.17 |
| CAD | 1773(51.4%) | 1536(47.8%) | 0.003 | 1031(48.7%) | 995(47.0%) | 0.27 |
| COPD | 908(26.3%) | 857(26.5%) | 0.87 | 575(27.2%) | 571(27.0%) | 0.89 |
| CKD | 120(3.5%) | 114(3.6%) | 0.88 | 83(3.9%) | 86(4.1%) | 0.81 |
| ASA class |  |  | <0.001 |  |  | 0.73 |
| 1-2 | 146(4.3%) | 513(17.0%) |  | 144(6.8%) | 180(8.5%) |  |
| 3 | 2243(65.6%) | 1871(62.1%) |  | 1489(70.4%) | 1461(69.1%) |  |
| 4-5 | 1031(30.1%) | 628(20.8%) |  | 482(22.8%) | 474(22.4%) |  |
| Ambulatory status |  |  | 0.18 |  |  |  |
| Full | 3257(95.3%) | 2788(94.5%) |  | 2003(94.7%) | 1997(94.4%) |  |
| Limited | >=11 | >=11 |  | >=11 | >=11 |  |
| Bedbound | <11 | <11 |  | <11 | <11 |  |
| Degree of stenosis on the treated side |  |  | <0.001 |  |  | 0.32 |
| 0-49% | 46(1.3%) | 89(2.7%) |  | 41(1.9%) | 56(2.6%) |  |
| 50-69% | 252(7.3%) | 230(7.1%) |  | 178(8.4%) | 185(8.7%) |  |
| 70-79% | 805(23.3%) | 700(21.6%) |  | 541(25.6%) | 539(25.5%) |  |
| 80-99% | 2181(63.2%) | 1628(50.3%) |  | 1264(59.8%) | 1228(58.1%) |  |
| Occluded | 30(0.9%) | 226(7.0%) |  | 30(1.4%) | 41(1.9%) |  |
| Missing | 136(3.9%) | 365(11.3%) |  | 61(2.9%) | 66(3.1%) |  |
| Prior medication |  |  |  |  |  |  |
| Anticoagulation | 511(14.8%) | 453(14.0%) | 0.35 | 307(14.5%) | 311(14.7%) | 0.86 |
| P2Y | 2956(85.7%) | 2285(70.6%) | <0.001 | 1691(80.0%) | 1642(77.6%) | 0.06 |
| Aspirin | 3080(89.3%) | 2713(83.9%) | <0.001 | 1858(87.8%) | 1837(86.9%) | 0.33 |
| Statin | 3067(88.9%) | 2532(78.3%) | <0.001 | 1795(84.9%) | 1771(83.7%) | 0.31 |
| History of radiation | 237(7.0%) | 276(8.9%) | 0.004 | 188(8.9%) | 194(9.2%) | 0.75 |
| Prior TIA on the treated side |  |  | <0.001 |  |  | 0.90 |
| No | 2832(82.1%) | 2485(76.7%) |  | 1688(79.8%) | 1686(79.7%) |  |
| Yes | 570(16.5%) | 545(16.8%) |  | 398(18.8%) | 403(19.1%) |  |
| Missing | 48(1.4%) | 208(6.4%) |  | 29(1.4%) | 26(1.2%) |  |
| Prior Stroke on the treated side |  |  | <0.001 |  |  | 0.21 |
| No | 2713(78.6%) | 2063(63.7%) |  | 1506(71.2%) | 1469(69.5%) |  |
| Yes | 722(20.9%) | 1158(35.8%) | <0.001 | 609 (28.8%) | 646 (30.5%) |  |
| Missing | 15(0.4%) | 17(0.5%) |  | <11 | <11 |  |

When comparing each of the stenting modalities with CEA, there was no significant difference in the risk of reintervention on comparison of patients undergoing TCAR vs. CEA (Table III). However, we did observe an increased risk of stroke in patients undergoing TCAR compared to CEA within six months (HR 1.07, 95% CI 1.07-1.77, p < 0.05) and an increased risk of mortality beyond six months (HR 1.46; 95% CI 1.15-1.81; p <0.05). Consistent with short-term outcomes studies, there was an increased risk of stroke and mortality in patients undergoing TFCAS compared to CEA which appears to be attributable at least in part to inferior perioperative outcomes (Table IV).

**Table III:** Risk of reintervention, stroke, and mortality with TCAR compared to CEA. Table IV: Risk of reintervention, stroke, and mortality of TFCAS compared to CEA. Table V: Multivariate analysis to identify predictors of reintervention.

|  | **HR** | **Lower level** | **Upper level** | **p-value** |
| --- | --- | --- | --- | --- |
| Reintervention |  |  |  |  |
| Within 6 months | 1.56 | 0.73 | 3.34 | 0.2563 |
| Beyond 6 months | 0.81 | 0.37 | 1.80 | 0.6048 |
| Stroke |  |  |  |  |
| Within 6 months | 1.38 | 1.07 | 1.77 | 0.0126 |
| Beyond 6 months | 1.21 | 0.82 | 1.77 | 0.3348 |
| Mortality |  |  |  |  |
| Within 6 months | 1.06 | 0.80 | 1.39 | 0.6943 |
| Beyond 6 months | 1.46 | 1.15 | 1.85 | 0.0019 |

**Table IV:**
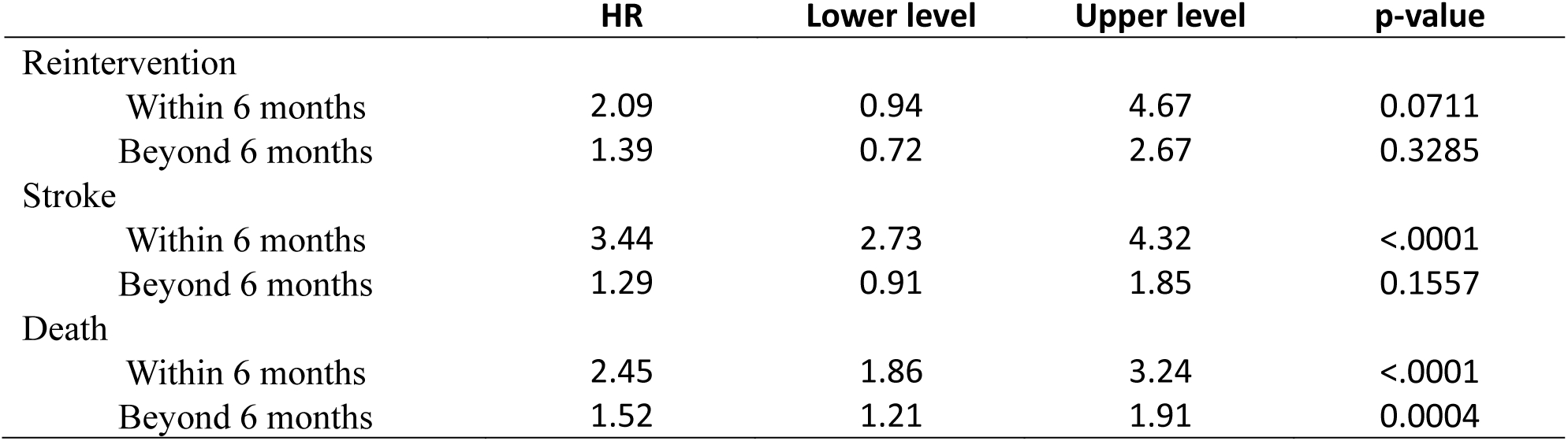
Risk of reintervention, stroke, and mortality of TFCAS compared to CEA.

**Table V:** Multivariate analysis to identify predictors of reintervention.

|  |  | **Hazard**<br>**Ratio** | **95% CI**<br>**lower limit** | **95% CI**<br>**upper limit** | **P value** |
| --- | --- | --- | --- | --- | --- |
| **Intervention (Ref CEA)** |  |  |  |  |  |
|  | **TFCAS** | 1.90 | 1.29 | 2.79 | 0.001 |
|  | **TCAR** | 1.09 | 0.70 | 1.69 | 0.710 |
| **Age** |  | 0.98 | 0.96 | 0.99 | 0.005 |
| **Female** |  | 1.24 | 0.97 | 1.58 | 0.086 |
| **Smoker** |  | 0.87 | 0.59 | 1.29 | 0.499 |
| **Prior Smoker** |  | 1.16 | 0.87 | 1.55 | 0.324 |
| **CAD** |  | 1.05 | 0.81 | 1.37 | 0.723 |
| **COPD** |  | 1.22 | 0.92 | 1.61 | 0.164 |
| **CKD** |  | 0.33 | 0.11 | 1.05 | 0.060 |
| **ASA (ref = 1,2)** |  |  |  |  |  |
|  | **ASA=3** | 1.09 | 0.65 | 1.81 | 0.749 |
|  | **ASA>4** | 1.25 | 0.72 | 2.18 | 0.424 |
| **Non-ambulatory** |  | 0.57 | 0.34 | 0.95 | 0.032 |
| **Degree of Stenosis** |  |  |  |  |  |
|  | **50-69%** | 1.23 | 0.42 | 3.59 | 0.710 |
|  | **70-79%** | 1.27 | 0.46 | 3.48 | 0.644 |
|  | **80-99%** | 1.50 | 0.55 | 4.04 | 0.426 |
|  | **Occluded** | 1.73 | 0.52 | 5.77 | 0.373 |
| **Anticoagulation** |  | 0.97 | 0.66 | 1.42 | 0.863 |
| **P2Y12** |  | 1.28 | 0.98 | 1.66 | 0.070 |
| **ASA** |  | 0.91 | 0.65 | 1.27 | 0.573 |
| **Statin** |  | 1.00 | 0.72 | 1.39 | 0.995 |
| **Radiation** |  | 1.67 | 0.99 | 2.79 | 0.053 |
| **TIA** |  | 1.04 | 0.69 | 1.57 | 0.866 |
| **Stroke** |  | 0.71 | 0.44 | 1.13 | 0.148 |

Finally, using multivariate logistic regression, we identified predictors of reintervention in patients undergoing carotid artery revascularization. TFCAS was independently associated with an increased risk of reintervention (HR: 1.90; 95% CI: 1.29-2.79, p < 0.001), while TCAR was not (Table V). Increased age was associated with a reduced risk of reintervention (HR: 0.98; 95% CI: 0.96-0.99; p < 0.01). Use of statins or antiplatelet agents, smoking status, or history of cerebrovascular accident, were not significantly associated with reintervention.

## Discussion

RCTs and large retrospective studies have consistently demonstrated that patients undergoing TCAR have improved perioperative outcomes compared to those undergoing TFCAS. However, the mid- to long-term outcomes following TCAR in the larger context of carotid revascularization remain poorly defined, specifically for reintervention rates. These will become particularly relevant in the coming years as TCAR gains more widespread use.^14^ In order to address this knowledge gap, we leveraged the VQI database linked to VISION Medicare claims data to assess outcomes following the three carotid revascularization approaches over a 36-month period. In this study, we found that patients undergoing CAS had increased risk of reintervention compared to CEA. Perhaps more importantly, we found that this signal was attributable to increased reintervention rates in patients undergoing TFCAS, as rates of reintervention between CEA and TCAR did not differ significantly. Controlling for preoperative and demographic factors, we found that patients who underwent TFCAS but not TCAR were at significantly increased risk of reintervention at 36 months.

Initial comparison of outcomes following all stenting modalities versus CEA revealed the expected findings of an increased risk of mortality and stroke that was primarily driven by inferior peri-operative outcomes. Our observation of increased mortality following stenting in the early post-operative period is consistent with the large retrospective analysis of over 80,000 patients by Columbo et al.,^15^ and is consistent with the results of the CREST trial.^16^ Somewhat surprisingly, we found that there was a significantly increased risk of reintervention within six months of the index operation but not beyond for patients undergoing CAS compared to CEA. This increased risk was apparent only on comparison of TFCAS to CEA but not TCAR to CEA. This could certainly be due in part to technical complications such as dissection, stent malposition and early thrombosis that would not be seen in open surgical reconstructions, but it is unclear why the same signal was not seen with TCAR. Restenosis in this time frame is often due to accelerated neointimal hyperplasia caused by endothelial damage and proliferation of the vascular smooth muscle cells in response to injury or technical complications.^17^ This pathophysiologic response may be greater in patients undergoing stenting than endarterectomy, leading to hemodynamically significant issues on early post-operative surveillance studies.

When comparing TCAR to TFCAS, we observed increased freedom from reintervention with separation of the curves at 6 months post-operatively. In our multivariate analysis, TFCAS increased the risk of re-intervention by nearly 2-fold. In fact, TFCAS, age, and ambulatory status were the only procedural or demographics variables that were associated with an increased risk of reintervention. Increased age and non-ambulatory status were associated with a moderately decreased risk of reintervention, possibly due to poor operative candidacy from comorbid conditions. While it may not be intuitive that different stenting modalities would have disparate longer term reintervention rates, it is important to note that TFCAS and TCAR are not simply the same procedure with delivery from distinct anatomic access points; differences in stent design could help explain the discrepancy.^18^ TCAR requires selective use of the Enroute conformable nitinol stent, whereas TFCAS procedures may use a variety of stents composed of alloys ranging from Elgiloy to nitinol.^19^ The Enroute stent is an open cell stent whereas transfemoral stenting can use multiple stent designs ranging from the closed cell Wallstent to the open cell Acculink. The differential effects of these stent materials or structure on the carotid endothelium have yet to be explored, but they may influence restenosis, measured clinically as increased reintervention rates.

CEA has long been the favored modality of carotid revascularization for both symptomatic and asymptomatic carotid stenosis. Peri-operative rates of stroke and death comparable to CEA have led to the Society of Vascular Surgery (SVS) to include TCAR along TFCAS as an option for asymptomatic carotid stenosis in patients with appropriate anatomy.^20^ Our observation that TFCAS but not TCAR is associated with increased risk of reintervention using propensity matched cohorts and real-world outcome data may further expand the use of TCAR as a favored stenting modality. The reduced rates of reintervention observed with TCAR provide further evidence that this stenting modality may be superior to TFCAS in the longer term, and on par with CEA with respect to risk of reintervention. Further study directed to assess outcomes of patients undergoing TCAR for symptomatic disease compared to asymptomatic disease could additionally provide support for use in asymptomatic patients. It is notable that in our study the presence of symptomatic carotid stenosis did not modify the risk of reintervention after revascularization.

This study has several limitations inherent to retrospective analyses. First, patients undergoing TCAR are required to have specific anatomic criteria. In the context of restenosis and reintervention, the most important of these requirements is that they have a 4-9 mm internal carotid artery (ICA). Narrow ICAs are more likely to have hemodynamically relevant restenosis, and patients with this anatomic risk factor are not considered for TCAR. In this retrospective study, we could not control for this degree of selection bias as anatomic parameters or imaging data were unavailable for analysis. Additionally, examination of our primary endpoint of reintervention and secondary endpoints of stroke and mortality was limited to a 36-month study period. Future studies that explore longer term reintervention rates may identify differences in reintervention rates between CEA and TCAR.

## Conclusions

Despite these limitations, we conclude that TFCAS portends an increased hazard of reintervention compared to TCAR and CEA in midterm outcomes. These data, along with previous perioperative safety data, support the use of TCAR as the favored stenting modality in patients with appropriate anatomy. Future RCTs performing head-to-head comparison of the two stenting modalities with longer term follow-up may further shed light on the mechanisms underlying our findings.

## Conflicts of Interest

The views expressed in this article are those of the authors and do not necessarily reflect the position or policy of the Department of Veterans Affairs or the United States government.

## Funding Information

No funding was provided.

## ARTICLE HIGHLIGHTS

### Key Findings

In matched cohorts of 4705 patients receiving either carotid endarterectomy or carotid stenting, we found that patients receiving carotid stenting had an increased risk of ipsilateral reintervention within 6 months of index procedure (HR 1.97; 95% CI: 1.11-3.50; p < 0.05). While no difference in ipsilateral reinterventions was observed between carotid endarterectomy or transcarotid artery revascularization (TCAR), patients undergoing transfemoral stenting had a higher risk of reintervention compared to TCAR (HR: 2.31; 95% CI: 1.05-5.11, p < 0.05).

### Take home Message

Amongst carotid revascularization modalities, carotid endarterectomy appears to be the most durable procedure. Amongst stenting modalities, TCAR is more durable than transfemoral stenting with a lower rate of reintervention.

### Table of Contents Summary

In a cohort of over 9000 patients, carotid endarterectomy was associated with reduced risk of ipsilateral reintervention compared to carotid artery stenting at 36 months follow-up. Carotid endarterectomy was the more durable revascularization approach amongst carotid revascularization modalities.

## Data Availability

The Vascular Quality Initiative datasets are national procedural datasets and the data within are available to institutions who participate in the Society for Vascular Surgery Quality Improvement Program after review board approval.

---

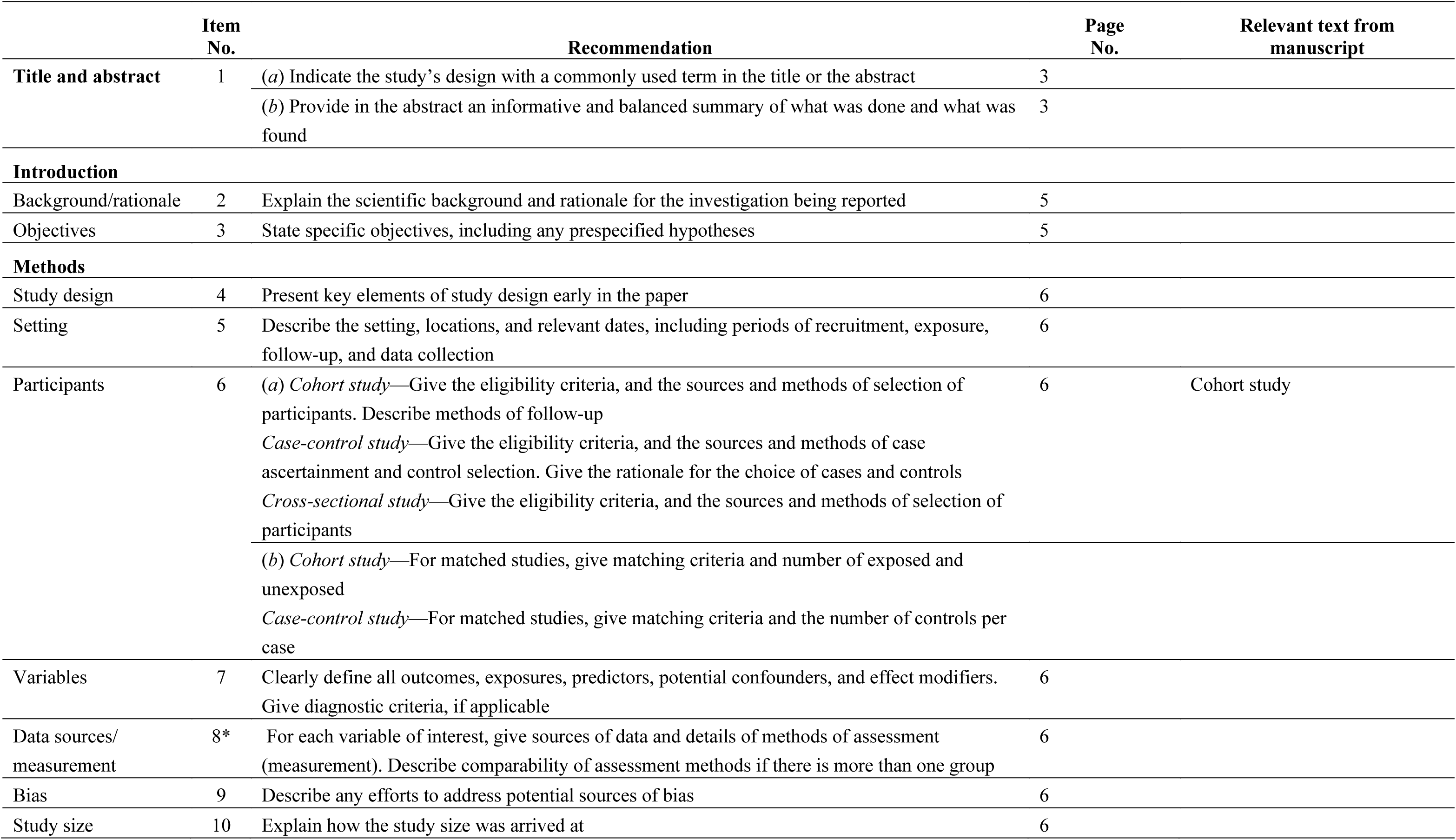
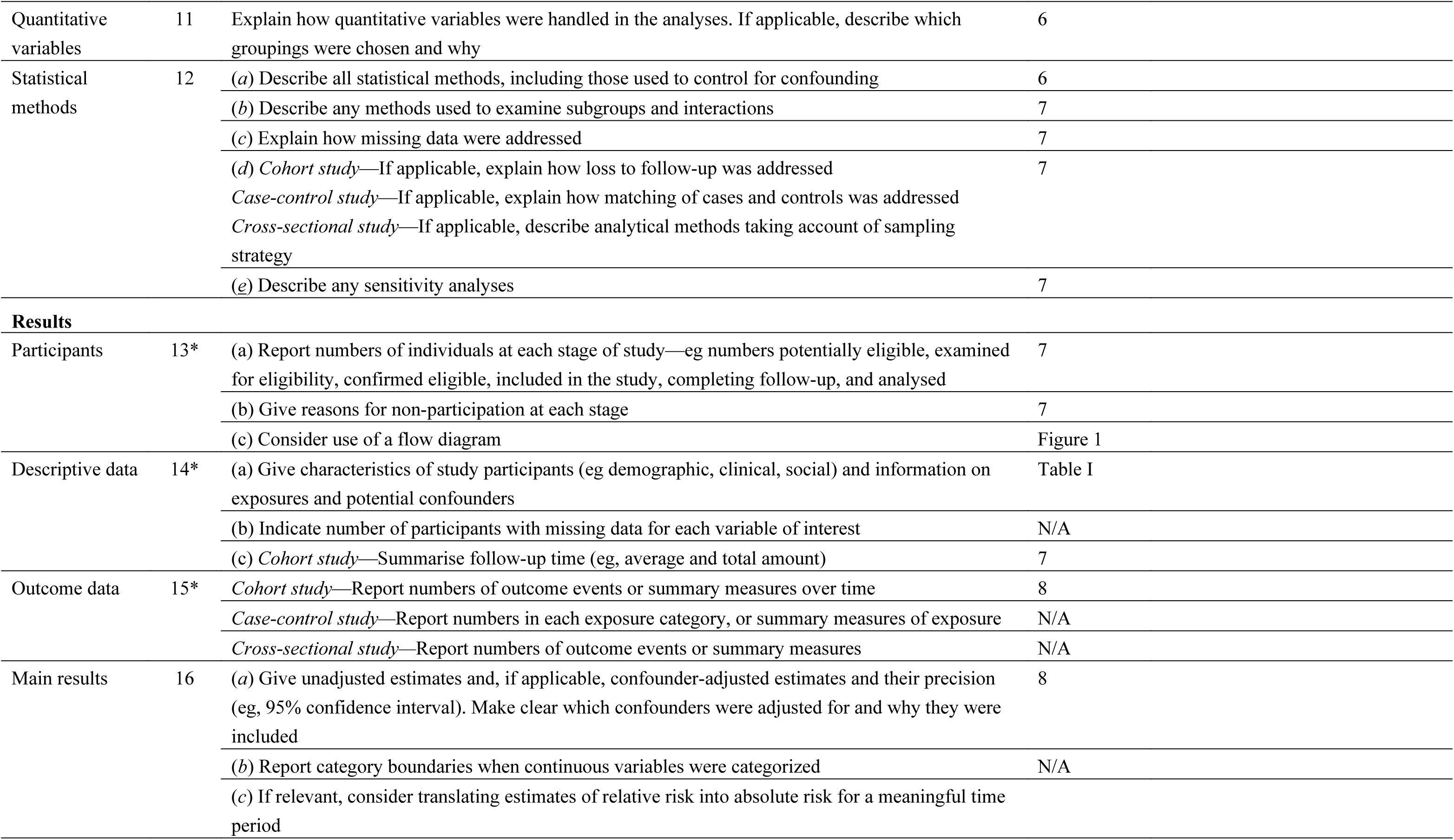
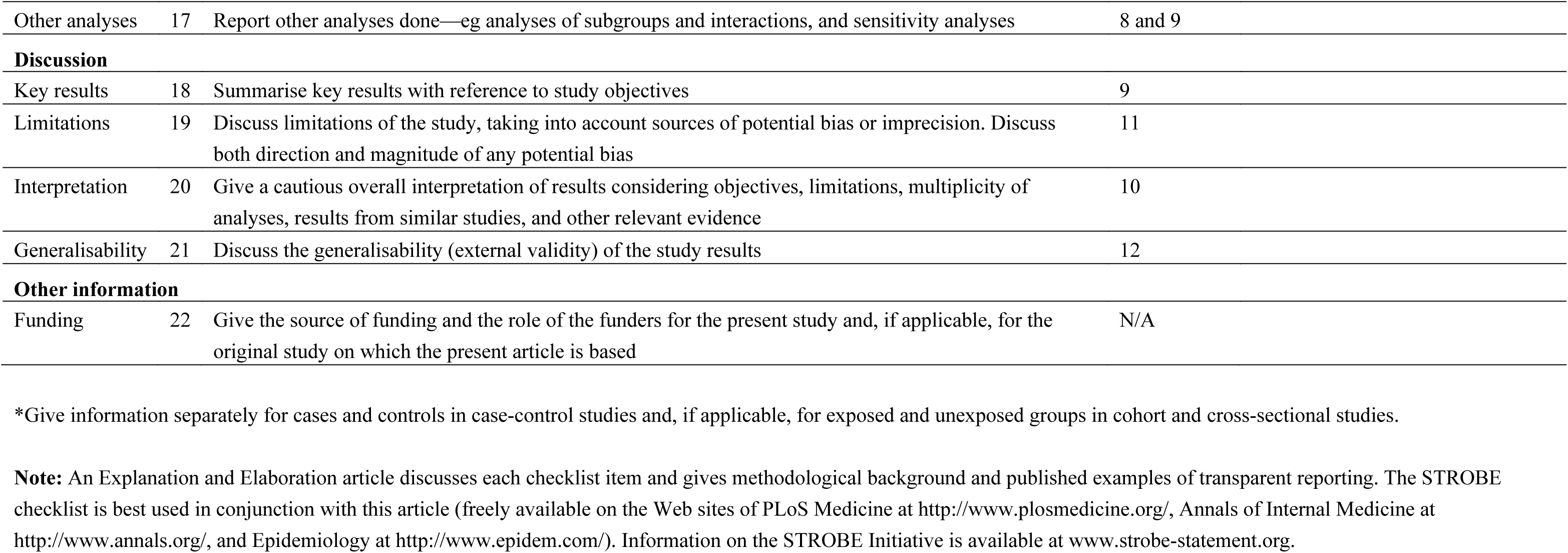
STROBE Statement–checklist of items that should be included in reports of observational studies.

|  | Item No. | Recommendation | Page No. | Relevant text from manuscript |
| --- | --- | --- | --- | --- |
| Title and abstract | 1 | (a) Indicate the study's design with a commonly used term in the title or the abstract | 3 |  |
|  |  | (b) Provide in the abstract an informative and balanced summary of what was done and what was found | 3 |  |
| **Introduction** |  |  |  |  |
| Background/rationale | 2 | Explain the scientific background and rationale for the investigation being reported | 5 |  |
| Objectives | 3 | State specific objectives, including any prespecified hypotheses | 5 |  |
| **Methods** |  |  |  |  |
| Study design | 4 | Present key elements of study design early in the paper | 6 |  |
| Setting | 5 | Describe the setting, locations, and relevant dates, including periods of recruitment, exposure, follow-up, and data collection | 6 |  |
| Participants | 6 | (a) *Cohort study*—Give the eligibility criteria, and the sources and methods of selection of participants. Describe methods of follow-up<br>*Case-control study*—Give the eligibility criteria, and the sources and methods of case ascertainment and control selection. Give the rationale for the choice of cases and controls<br>*Cross-sectional study*—Give the eligibility criteria, and the sources and methods of selection of participants<br>(b) *Cohort study*—For matched studies, give matching criteria and number of exposed and unexposed<br>*Case-control study*—For matched studies, give matching criteria and the number of controls per case | 6 | Cohort study |
| Variables | 7 | Clearly define all outcomes, exposures, predictors, potential confounders, and effect modifiers. Give diagnostic criteria, if applicable | 6 |  |
| Data sources/measurement | 8* | For each variable of interest, give sources of data and details of methods of assessment (measurement). Describe comparability of assessment methods if there is more than one group | 6 |  |
| Bias | 9 | Describe any efforts to address potential sources of bias | 6 |  |
| Study size | 10 | Explain how the study size was arrived at | 6 |  |

|  |  |  |  |
| --- | --- | --- | --- |
| Quantitative variables | 11 | Explain how quantitative variables were handled in the analyses. If applicable, describe which groupings were chosen and why | 6 |
| Statistical methods | 12 | (a) Describe all statistical methods, including those used to control for confounding | 6 |
|  |  | (b) Describe any methods used to examine subgroups and interactions | 7 |
|  |  | (c) Explain how missing data were addressed | 7 |
|  |  | (d) *Cohort study*—If applicable, explain how loss to follow-up was addressed | 7 |
|  |  | *Case-control study*—If applicable, explain how matching of cases and controls was addressed |  |
|  |  | *Cross-sectional study*—If applicable, describe analytical methods taking account of sampling strategy |  |
|  |  | (e) Describe any sensitivity analyses | 7 |
| **Results** |  |  |  |
| Participants | 13* | (a) Report numbers of individuals at each stage of study—eg numbers potentially eligible, examined for eligibility, confirmed eligible, included in the study, completing follow-up, and analysed | 7 |
|  |  | (b) Give reasons for non-participation at each stage | 7 |
|  |  | (c) Consider use of a flow diagram | Figure 1 |
| Descriptive data | 14* | (a) Give characteristics of study participants (eg demographic, clinical, social) and information on exposures and potential confounders | Table I |
|  |  | (b) Indicate number of participants with missing data for each variable of interest | N/A |
|  |  | (c) *Cohort study*—Summarise follow-up time (eg, average and total amount) | 7 |
| Outcome data | 15* | *Cohort study*—Report numbers of outcome events or summary measures over time | 8 |
|  |  | *Case-control study*—Report numbers in each exposure category, or summary measures of exposure | N/A |
|  |  | *Cross-sectional study*—Report numbers of outcome events or summary measures | N/A |
| Main results | 16 | (a) Give unadjusted estimates and, if applicable, confounder-adjusted estimates and their precision (eg, 95% confidence interval). Make clear which confounders were adjusted for and why they were included | 8 |
|  |  | (b) Report category boundaries when continuous variables were categorized | N/A |
|  |  | (c) If relevant, consider translating estimates of relative risk into absolute risk for a meaningful time period |  |

|  |  |  |  |
| --- | --- | --- | --- |
| Other analyses | 17 | Report other analyses done—eg analyses of subgroups and interactions, and sensitivity analyses | 8 and 9 |
| **Discussion** |  |  |  |
| Key results | 18 | Summarise key results with reference to study objectives | 9 |
| Limitations | 19 | Discuss limitations of the study, taking into account sources of potential bias or imprecision. Discuss both direction and magnitude of any potential bias | 11 |
| Interpretation | 20 | Give a cautious overall interpretation of results considering objectives, limitations, multiplicity of analyses, results from similar studies, and other relevant evidence | 10 |
| Generalisability | 21 | Discuss the generalisability (external validity) of the study results | 12 |
| **Other information** |  |  |  |
| Funding | 22 | Give the source of funding and the role of the funders for the present study and, if applicable, for the original study on which the present article is based | N/A |

\*Give information separately for cases and controls in case-control studies and, if applicable, for exposed and unexposed groups in cohort and cross-sectional studies.
**Note:** An Explanation and Elaboration article discusses each checklist item and gives methodological background and published examples of transparent reporting. The STROBE checklist is best used in conjunction with this article (freely available on the Web sites of PLoS Medicine at http://www.plosmedicine.org/, Annals of Internal Medicine at http://www.annals.org/, and Epidemiology at http://www.epidem.com/). Information on the STROBE Initiative is available at www.strobe-statement.org.

## Notes

### Competing Interest Statement

The authors have declared no competing interest.

### Funding Statement

No external funding was received.

### Author Declarations

Stanford University IRB-57282

